# Clinical characteristics of COVID-19 patients admitted to Intensive Care Unit in Panama during the first pandemic wave admissions in 2020

**DOI:** 10.1101/2022.05.08.22274803

**Authors:** Silvio Vega, Artur Gomez Blizniak, Simón Theoktisto Ballanis, Gabriel Cisneros, David Villalobos, Ana Kathleen Armuelles, Luis Moreno, Francis Fusumada, Johanna Gonzalez, Emilio Romero, Ivonne Torres-Atencio, Idalina Cubilla-Batista, Amador Goodridge

**Affiliations:** Complejo Hospitalario Metropolitano Dr. Arnulfo Arias Madrid, Caja de Seguro Social, Panamá, PANAMA; Departamento de Fisiología y Comportamiento Animal, Facultad de Ciencias Naturales, Exactas y Tecnología, Universidad de Panamá, Panamá, PANAMA; Departamento de Farmacología, Facultad de Medicina, Universidad de Panamá, Panamá, PANAMA; Centro de Biología Celular y Molecular de las Enfermedades, Instituto de Investigaciones Científicas y Servicios de Alta Tecnología (INDICASAT-AIP), City of Knowledge, PANAMA; Hospital Rafael Estevez, Caja de Seguro Social, Aguadulce, Coclé, PANAMA

**Keywords:** Intensive Care Unit, COVID-19, Clinical characteristics

## Abstract

The severe acute respiratory syndrome Coronavirus 2 (SARS COV-2) caused a global pandemic of COVID-19. Most of people affected are admitted to hospital with various grades of ADRS. A small proportion of these patients requires intensive care unit management and treatment. However not all of them survive. This study aims to describe the epidemiological and clinical characteristics of patients admitted to the intensive care units in Panama main hospital in the first six months of pandemic with available information. Special focus has been oriented to blood and respiratory biomarkers to correlate with survivors and non-survivors. Our results show that patients between 56-75 years old, with hypertension, obesity, and diabetes comorbid conditions are more likely to die in intensive care units. Regarding the PaFi ratio, we observed a greater proportion of non-survivor with values less than 200. The triglycerides, urea nitrogen, creatinine and procalcitonin levels resulted significantly higher in those non survivors. During clinical management, half of patient that were administered Tocilizumab did not survived. These results support the notion that age, comorbidities as well as therapeutic management of patient in intensive care units contribute to the final outcome. We recommend reinforcing patient care strategy, especially in those patients with clinical conditions that favor fatal outcomes.

## INTRODUCTION

The severe coronavirus disease by SARS-CoV-2 (COVID-19) outbreak emerged in Wuhan, China in December 2019. In late January 2020, the World Health Organization (WHO) declared it the sixth international public health emergency due to its rapid expansion. After more than 200,000 cases reported in a short time, in early March the WHO declared it a global pandemic [1]. Today there are more than 271 million confirmed cases, causing just over 5.3 million deaths: with the highest report of active cases in the United States, India, Brazil, United Kingdom, and Russia (**Figure 1**). In Central America, Panama reported the first case of COVID-19 on March 8 and to date we have reached 735,000 cases, with a more than of 7,950 deaths, with an affectation of 11,075 cases per 100,00 inhabitants and 170.2 deaths per 100,000 inhabitants: the fourth highest in the Americas region and the 36^th^ position among the countries reporting to the WHO.

**Figure 1.**
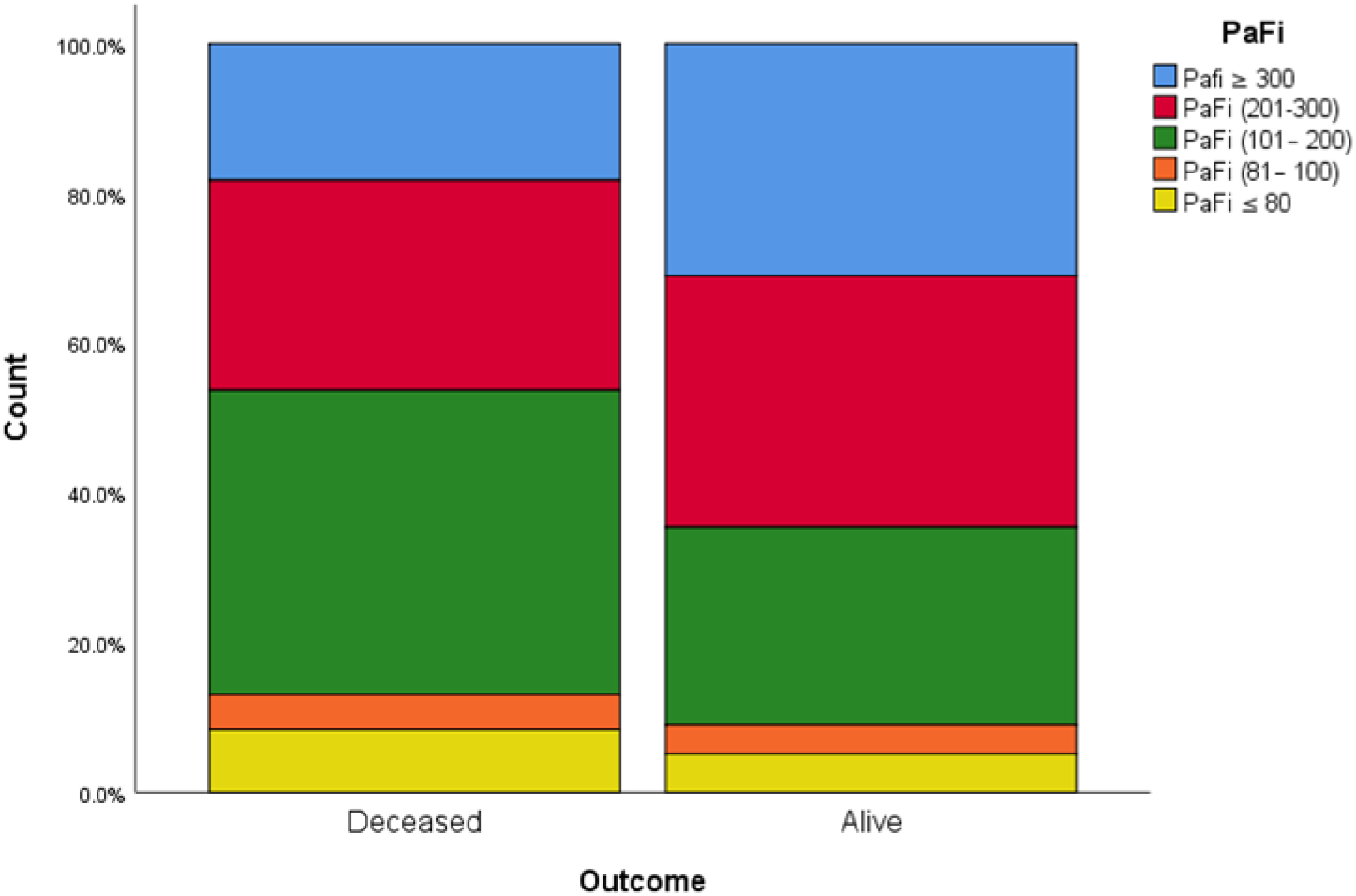

The clinical syndrome COVID-19 is caused by a new coronavirus called SARS-CoV-2 (severe acute respiratory syndrome coronavirus 2) and the new variants have been discovered since de first strain reported [2] (Imai et al., 2021; Khan et al., 2021; Tang et al., 2021; Zhou et al., 2020) (Zhou P, et al). This beta-coronavirus has an envelope that covers a nucleocapsid that surrounds a single-stranded positive-sense RNA. SARS-CoV2 has been shown to be highly efficient in its transmission from person to person. After entering the respiratory system, SARS-CoV2 induces acute respiratory distress syndrome (ARDS), a serious disease characterized by interstitial pneumonia and the rapid development of acute respiratory distress syndrome, septic shock with high levels of reactive species of acute phase and characteristics of macrophage activation syndrome. Briefly, SARS-CoV-2 enters the cell using as a receptor the angiotensin converting enzyme 2 (ACE-2), a membrane exopeptidase presents mainly in the kidney, endothelium, lungs and the heart [3]. The ACE 2 function is the transformation of Angiotensin I into Angiotensin 1-9 and Angiotensin II into Angiotensin 1-7. These products have vasodilatory, antifibrosis, ant inflammatory effects and promote natriuresis. They are all effects, therefore, that reduce blood pressure, against regulating the action of Angiotensin II. ACE2 has been linked to protection against hypertension, arteriosclerosis, and other vascular and pulmonary processes. Unfortunately, in severe cases of ARDS due to COVID-19, very high levels of Angiotensin II are present, highly correlated with the SARS-CoV-2 viral load and lung damage. This imbalance of the renin-angiotensin-aldosterone system could be related to the inhibition of ACE2 by the virus and consequently the systemic failure that induces death [4].

During the health crisis of COVID-19, the differentiation of seriously ill patients in ARDS is critical for the decision-making of admission and management in intensive care units (ICU). In Panama, cases increased between 25% and 45% daily during the first peak of epidemic. From the experiences of China, Italy, and Spain, we know that up to 20% of positive COVID-19 cases will require hospital care and of this group, 1 in 4 will be severe and require ICU management [5] (Mahase, 2020). At this point in the pandemic, much more is known about the characteristics of the patients, and it is suspected that some of this data may help to predict or anticipate outcomes.

The studies that were carried out in countries such as China, the United States, Italy and Peru found that of the patients who enter ICUs between 68-85% have at least one comorbidity, in addition, the mean age of ICU admission found in different studies has ranged from 53 to 73 years, that being older than 75 years, having a BMI> 40 and being a man are the risk factors associated with severe disease[6-9]. In Panama, COVID 19 patient 0 was admitted at the beginning of March for suspected *Mycoplasma pneumoniae* pneumonia, but the COVID-19 diagnostic confirmation was obtained one week later, in post-mortem examination. However, the clinical characteristics of patients admitted to ICU in Panama have not been described yet. Our study aims to conduct an evaluation of the epidemiological and clinical characteristics of patients with COVID-19 admitted to the ICU at the main COVID-19 hospital in Panama during April to September 2020 period (the first six months of admissions in the ICU Dr. Arnulfo Arias Madrid Complex Hospital in Panamá-CHMDrAAM, first wave of our pandemic period with the information we had at that time).

## METHODOLOGY

### Study site and participants

This study was conducted between April and September 2020 in the Hospital Dr. Arnulfo Arias Madrid (CHMDrAAM) located in Panama City, Republic of Panama. This is a third level hospital for adults with 830 beds. This hospital was transformed into the main COVID-19 hospital in our country, including ICU, Respiratory Specials Care Unit, Intermediate Care Unit, during the highest pandemic peak. A total of 553 medical files from ICU patients were reviewed. The data was collected by two practitioners who visited the ICU daily and reviewed the records of each patient. This study was approved by the National Bioethics Committee Board EC-CNBI-2020-10-103.

### Data collection

This was an observational, descriptive cross-sectional study. For this study we reviewed the records of patients diagnosed with ARDS by COVID-19 and admitted to any of the intensive care units of the Dr. Arnulfo Arias Madrid Hospital Complex during study period. Briefly, each week, the information of the patients admitted to any of the authorized ICUs was annotated. For the collection of the individual’s data, the medical records were reviewed directly from the clinical record using the data collection tool designed for this purpose. All the records of patients admitted from April to September reviewed twice a week while they were hospitalized in an ICU ward. In addition, other data records such as notes, and books or nursing records were reviewed.

### Statistical analysis

We determined the total and frequencies for all study variables. Patient information was analyzed, such as demographics, disease onset before admission to the ICU, history of previous treatment and medication for other diseases, history of previous treatment, and medication for other diseases, among other clinical aspects. All statistical testing will be carried out in SPSS v27 for Windows.

## RESULTS

The first COVID-19 patient at the ICU of this center was admitted on March 3, 2020. At the end of September, a total of 553 patients were admitted with a diagnosis of Covid-19, of which 322 (58.2%) men and 231 (41.8%) women. All patients had upon admission a positive SARS-CoV2 RT-PCR test. The age ranged from 16 to 87 years, average 62.

The range of days of stay in the ICU was 1-71, with a mean stay of 16, 17 days for survivors and 14 days for no survivors. **Table 1**, summarizes the age, sex and age groups distribution. The most affected age group were between 56 and 75 years (55%, 303 patients), where we also found the highest mortality (63%, 204 patients). The overall mortality was 58.9%, where women had 39.6% and men 60.4%.

**Table 1:**
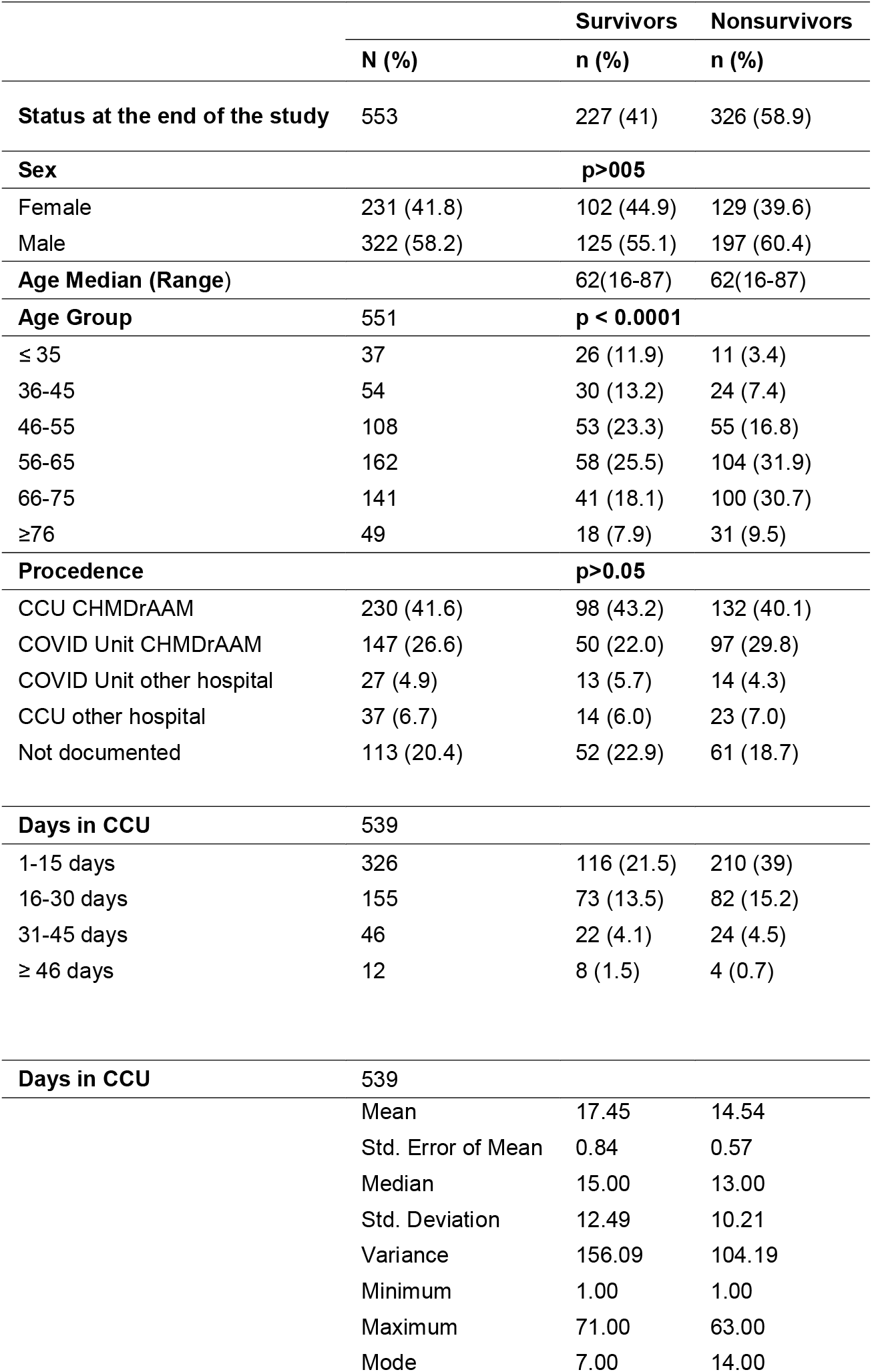
Demographical characteristic of ICU admitted patients March – September 2021

Before admission to the ICU, 41.6% of the patients were previously at the Emergency room at the CHMDrAAM, 26.6% were hospitalized in a COVID-19 ward at the CHMDrAAM, or 11.6% directly in ICU COVID-19 ward from other hospital mostly from the metropolitan area. Although it was impossible to collect all the information for each case (due to missing data, often due to the absence of a family member who could provide us the information or because the patient was not in a clinical condition to answer the questions), among the reported comorbidities, the most prevalent were cardiovascular diseases with 245 (45.7%) subjects from whom 214 (88%) had hypertension. Other comorbidities are listed in **Table 2**, where the presence of hypertension, diabetes mellitus and obesity stand out, as the main three. High mortality was observed in the group of diabetic’s patients, where 71(59.6%) died. Patients who were admitted with bronchial asthma rapidly worsened and 50% of them died. Two patients admitted with systemic lupus erythematosus and 4 patients with acute renal failure, also died.

**Table 2:** Comorbidities and medication associated among COVID-19patients admitted to ICU (subgroup)

| Comorbidities (n= total number reported) |  | Survivors<br>n (%) | Nonsurvivors<br>n (%) |
| --- | --- | --- | --- |
| Hypertension | 214 | 90 (42.1) | 124 (57.9) |
| Diabetes Mellitus 2 | 119 | 48(40.4) | 71 (59.6) |
| Obesity | 63 | 23 (13.7) | 40 (23.9) |
| Asthma | 42 | 21 (50) | 21 (50) |
| Chronic Kidney disease | 22 | 9 (40.9) | 13 (59.1) |
| Ischemic heart | 13 | 2 (15.4) | 11 (84.6) |
| Hematological Disorders | 11 | 2 | 9 |
| Other Cardiovasc . diseases | 9 | 4 (45.5) | 5 (55.5) |
| EPOC | 5 | 1 (20) | 4 (80) |
| Arrhythmias | 4 | 2 (50) | 2 (50) |
| Acute Kidney disease | 4 | 0 | 4 (100) |
| Dyslipidemia | 2 | 1 (50) | 1 (50) |
| Neoplasia | 6 | 1 (16.7) | 5 (83.3) |
| Hepatic cirrhosis | 1 | 0 | 1 (100) |
| Previous ECV | 5 | 3 (60) | 2 (40) |
| Other Rheumatoid diseases | 5 | 3 (60) | 2 (40) |
| Rheumatoid arthritis | 6 | 3 (50) | 3 (50) |
| Lupus | 2 | 2 (100) | 0 |
| Epilepsies | 3 | 1 (33.3) | 2 (66.6) |
| **Non-pathological antecedents** |  |  |  |
| Smoke | 16 | 6 (4.6) | 10 (7.6) |
| Alcohol consumption | 22 | 12 (22.2) | 10 (12.7) |
| **Chronic Use Medications Reported** |  |  |  |
| IECA | 29 | 13 (5.7) | 16 (4.9) |
| ARA2 | 35 | 16 (7.0) | 19 (5.8) |
| Beta-blockers | 17 | 6 (2.6) | 11 (3.3) |
| Inhaled corticosteroids |  |  |  |
| Oral corticosteroids | 8 | 5 (2.2) | 3 (1.0) |
| Statins | 26 | 12 (5.3) | 14 (4.3) |
| Diuretics | 38 | 16 (7.0) | 22 (6.7) |
| Ca Antagonists | 78 | 31 (13.7) | 47 (14.4) |
| Oral hypoglycemic | 55 | 21 (9.3) | 34 (10.4) |
| Insulin | 16 | 8 (3.5) | 8 (2.4) |

**Table 3** presents the imaging studies performed on the patients as well as some rigorous invasive procedures. All patients were admitted with a chest X-ray with compatible images of acute lung disease due to COVID-19. Of approximately 700 critical care ultrasounds performed on patients with COVID-19 in Panama by the Critical Care Ultrasound Unit (CCUU) leaded by Dr. Gómez, 345 were performed during the first six months of the Pandemic at the CHMDrAAM, of these, 320 were critical care echocardiograms, 21 lung ultrasounds, 3 transcranial Doppler, 1 fast abdominal ultrasound. Chest CT were performed in 3 patients (0.7%) and Chest angio-CT only in 6 patients (1.1%).

**Table 3:**
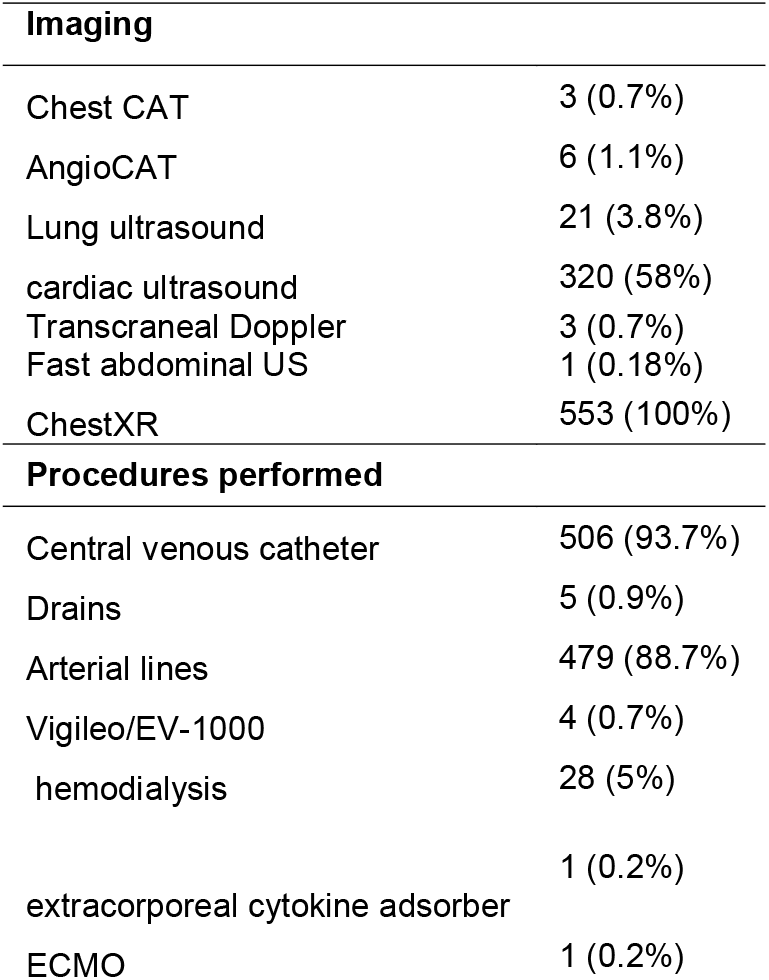
Diagnosis tool among COVID-19patients admitted to ICU and invasive procedures:

The main admission diagnoses were pneumonia (96%). All patients required endotracheal intubation upon admission due to the presence of acute respiratory failure indicated by increased respiratory rate and hypoxemia, as well as other laboratory variables detailed in **Table 3**. Initial COVID-19 pneumonia, in the majority of patients 531 (96.4%) progressed to ARDS.

The most frequent causes of death in the ICU of the patients in our study were: Multiorgan dysfunction syndrome, followed by acute respiratory distress syndrome secondary to COVID-19 pneumonia, shock of different etiologies (septic, obstructive, cardiogenic and mixed), acute renal failure, refractory pneumothorax complicated by bronchopleural fistula. **Table 4**.

**Table 4:** Diagnostics in ICU.

|  | N | Survivors | Nonsurvivors |
| --- | --- | --- | --- |
| **Pneumonia** | 531 (96%) | 219 (41.3%) | 312 (58.7%) |
| **Congestive heart failure** | 4 | 2 (50%) | 2 (50%) |
| **Acute coronary syndrome** | 3 | 1 (33.3%) | 2 (66.6%) |
| **Acute Respiratory Insufficiency** | 12 | 7 (58.4%) | 5 (41.6%) |
| **Acute Renal Failure** | 48 | 11 (22.9%) | 37 (77%) |
| **Hemophagocytic Syndrome** | 2 | 2 (100%) | 0 |
| **Pulmonary thromboembolism** | 1 | 0 | 1 (100%) |
| **Multiorgan dysfunction syndrome** | 9 | 0 | 9 (100%) |
| **Nosocomial infection** | 8 | 2 (25%) | 6 (75%) |
| **TBC** | 4 | 3 (75%) | 1 (25%) |
| **HIV** | 5 |  | 5 (100%) |
| **Shock** | 98 | 33 (33.6%) | 65(69.4%) |
| **Cardiogenic** | 1 | 1(1%) | 0 |
| **Distributive** | 87 | 30 (30.6%) | 57 (58.1%) |
| **Mixed .** | 10 | 2 (2%) | 8 (8.2%) |

### Therapeutic approach

Upon admission to the ICU, all patients had received treatment with some antimicrobial, such as azithromycin (79%), hydroxychloroquine (70%) and ivermectin (67%). These three drugs were included in a government authorized kit for early use in COVID-19. Other antibiotics used before admission were ceftriaxone (58%), erythromycin (52%), ampicillin (32%), and ciprofloxacin (28%).

All patients received corticosteroids upon admission, according to local guidelines (some received hydrocortisone, 200 mg daily in case of septic shock, others received dexamethasone 6-10 mg daily. Others received initial bolus prednisone 250 mg, followed by doses of 40-60 mg twice daily for 7-10 days. **Table 5** summarizes the medications used in the treatment of the patients. Briefly, 62% of the patients required vasopressors, of which the 51% (275 patients) was norepinephrine. The use of inotropic medications was supported by hemodynamic measurements and echocardiography and was required by 20% of the patients; levosimendan, dobutamine, and milrinone were used. 86% of the patients received anticoagulants, heparin 8% (42) and enoxaparin 78.2% (442). Heparin was used more in the patient with kidney failure. Three biotechnological drugs were included in the treatment. Tocilizumab, in patients with large elevations of IL-6; immunoglobulins and human immunoglobulins enriched with IgM, IgA. Antiparasitic drugs were used at the beginning of the pandemic, also 56% of the patients received hydroxychloroquine (800 mg daily). However, its use began to be reduced due to the presentation of arrhythmias in 22% of the patients (supraventricular tachycardia, atrial arrhythmias, and QT segment prolongation). Ivermectin was used in only 12% of the patients. In none of the patients treated with these drugs we observed benefits inherent to their use.

**Table 5:** Medication applied to COVID-19patients admitted to ICU

|  | n | Survivors | Nonsurvivors |
| --- | --- | --- | --- |
| **Corticosteroids** |  |  |  |
| Hydrocortisone | 90 | 24 (26.7) | 66 (73.3) |
| Dexametasone | 314 | 121 (38.6) | 193 (61.4) |
| Prednisone | 150 | 67 (44.7) | 83 (55.3) |
| **Vasopressors** |  |  |  |
| Norepinephrine | 275 | 115 (42.2) | 158 (57.8) |
| Vasopresin | 62 | 12 (19.4) | 50 (80.6) |
| **Inotropics** |  |  |  |
| Levosimendan | 24 | 6 (25.) | 18 (75) |
| Milrinone | 32 | 12 (37.5) | 20 (62.5) |
| Dobutamine | 56 | 23 (41.1) | 33 (58.9) |
| **Anticoagulants** |  |  |  |
| Heparin | 42 | 15 (35.8) | 27 (64.2) |
| Enoxaparin | 422 | 191 (45.3) | 231 (54.7) |
| Antiretrovirals |  |  |  |
| Remdesivir | 7 | 2 (28.6) | 5 (71.4) |
| Lopinar/Ritonavir | 18 | 12 (66.6) | 6 (33.3) |
| **Biologics** |  |  |  |
| Tocilizumab | 96 | 45 (46.9) | 51 (53.1) |
| Immunoglobulins | 153 | 63 (41.2) | 90 (58.8) |
| Pentaglobin | 6 | 2 (33.3) | 4 (66.6) |
| **Antiparasitic** |  |  |  |
| Ivermectin | 55 | 27 (49.1) | 28 (50.9) |
| Hydroxychloroquine | 63 | 33 (52.4) | 30 (47.6) |
| **Antibiotics** |  |  |  |
| Cephalosporin | 256 | 106 (42.2) | 148 (57.8) |
| Azithromycin | 228 | 103 (45.7) | 124 (54.3) |
| Carbapenemics | 198 | 83 (42) | 115 (58) |
| Linezolid | 39 | 10 (25.7) | 29 (74.3) |
| Levofloxacin | 93 | 52 (66) | 41 (44) |
| Vancomycin | 252 | 103 (42.2) | 148 (57.8) |
| Tazocin | 178 | 75 (42.7) | 102 (57.3) |
| TMP_SMX | 19 | 13 (68.5) | 6 (31.5) |
| Ciprofloxacin | 16 | 8 (56.3) | 7 (43.7) |
| Teicoplanin | 8 | 4 (50) | 4 (50) |
| Cefepime | 93 | 38 (42) | 54 (58) |
| Aminoglycosides | 48 | 23 (48) | 25 (52) |
| **Antifungals** |  |  |  |
| Amphotericin | 6 | 5 (83.4) | 1 (16.6) |
| Fluconazole | 102 | 40 (39.3) | 62 (60.7) |
| Echinocandins | 48 | 17 (35.5) | 31 (64.5) |

### Disease progression biomarkers

Blood biomarkers to monitor COVID-19 progression among ICU patients resulted diverse. Blood biomarker upon admission to ICU including glycemia, urea nitrogen, creatinine as well as C-reactive protein (CRP) D-dimer, ferritin, Interleukin 6, troponin and procalcitonin and others are shown in **table 6**. For example, we noted that COVID-19 patients withhold an average CRP level of 123.97 UI (SD 93.41 UI). Whereas Procalcitonin was elevated in most patients admitted to ICU (mean 4.43, range 0.02 - 160). Only in a few cases (30%) could it be correlated with the presence of concomitant bacterial infection. Regarding the blood cell parameters, we observed that 69% of ICU newly admitted patients had lymphocyte-predominant leukocytosis. When comparing the blood biomarkers between survivors and non survivors, we noted an overall increase of blood biomarker levels. **Table 7** show the different blood biomarker level between survivors and non-survivors after ICU admission.

**Table 6:** Initial Laboratory Results at the ICU.

|  | n | Mean | SD | Median | Range |
| --- | --- | --- | --- | --- | --- |
| PCR | 119 | 123.97 | 93.41 | 103.98 | 0.10-326 |
| Procalcitonin | 109 | 4.43 | 17.05 | 0.24 | 0.02-160 |
| Troponins | 24 | 143.2 | 415.06 | 0.2 | 0.00-1918 |
| CPK | 250 | 412.37 | 1034.9 | 141.5 | 12.00-12795 |
| CK_MB | 101 | 60.07 | 143.89 | 16 | 1.20-714 |
| LDH | 91 | 547.31 | 511.26 | 403 | 148.00-3429 |
| Dimer D | 144 | 4529.51 | 17315.25 | 943.86 | 1.80-197250 |
| Creatinine | 372 | 1.63 | 2.06 | 0.94 | 0.30-20 |
| Nitrogen_urea | 364 | 39.23 | 31.13 | 30.1 | 4.70-214 |
| AST_2 | 343 | 87.52 | 445.33 | 39 | 8.00-7182 |
| ALT_2 | 344 | 88.22 | 300.67 | 50.5 | 4.00-5431 |
| Albumin | 371 | 3.06 | 1.28 | 3 | 1.40-25 |
| Total Bilirubins | 351 | 0.76 | 1.08 | 0.5 | 0.10-10 |
| Direct bilirubin | 370 | 0.35 | 0.69 | 0.2 | 0.00-10 |
| Indirect Bilirubin | 344 | 0.3 | 0.26 | 0.27 | 0.00-3 |
| Ferritin 2 | 139 | 925.25 | 460.34 | 902 | 75.10-1821 |
| Vitamin D | 1 | 1500 |  | 1500 | 1500.00-1500 |
| Triglycerides | 327 | 207.9 | 116.05 | 184 | 45.00-936 |
| IL_6_2 | 137 | 248.06 | 413.88 | 70.69 | 1.15-1620 |
| BNP_2 | 5 | 1490.4 | 1465.24 | 866 | 240.00-3857 |
| TP | 166 | 13.02 | 1.94 | 12.75 | 9.50-20 |
| TPT | 166 | 33.58 | 16.4 | 32.1 | 11.40-192 |
| INR | 151 | 1.14 | 0.17 | 1.12 | 0.84-1 |
| Fibrinogen | 89 | 438.61 | 151.81 | 471 | 64.00-721 |
| Thromboelastography | 0 |  |  |  | 0.06-149 |
| Leukocytes | 397 | 14.61 | 10.36 | 12.5 | 0.00-218 |
| Lymphocytes | 391 | 7.86 | 13.59 | 5.49 | 1.76-862 |
| Neutrophils | 392 | 88.89 | 41.07 | 89.7 | 253.00-377 |
| Lymph / leukocytes | 2 | 315 | 87.68 | 315 | 10.20-642 |
| Platelets | 389 | 246.02 | 111.13 | 241 |  |

**TABLE 7.** **Differences between survivors and deceased observed in selected laboratory results at ICU admission**

|  | n | Values | Survivors | Non-survivors | p value |
| --- | --- | --- | --- | --- | --- |
|  |  |  | n (%) | n (%) |  |
| PCR >5 | 119 | < 5 | 2 (1.7) | 6 (5) | 0.32 |
|  |  | >5 | 49 (41.9) | 62 (52.1) |  |
| PCT >2 | 108 | <2 | 42 (38.9) | 44 (40.7) | **0.03** |
|  |  | >2 | 5 (4.6) | 17 (15.7) |  |
| Dimer D > 500 | 144 | <500 | 13 (9) | 12 (8.3) | 0.23 |
|  |  | > 500 | 46 (31.9) | 73 (50.7) |  |
| Creat >1.18 | 372 | < 1.18 | 123 (33.1) | 120 (32.3) | **0.001** |
|  |  | > 1.18 | 35 (9.4) | 94 (25.3) |  |
| BUN >20 | 360 | < 20 | 55 (15.3) | 38 (10.6) | **0.001** |
|  |  | > 20 | 97 (26.9) | 170 (47.2) |  |
| Ferritine >336 | 138 | < 336 | 7 (5.1) | 7 (5.1) | 0.39 |
|  |  | > 336 | 47 (34.1) | 77 (55.8) |  |
| Trigl>150 | 324 | < 150 | 61 (18.8) | 56 (17.3) | **0.02** |
|  |  | > 150 | 80 (24.7) | 127 (39.2) |  |
| IL-6>6.4 | 128 | < 6.4 | 5 (3.9) | 3 (2.3) | 0.15 |
|  |  | > 6.4 | 45 (35.2) | 83 (64.8) |  |
| Leukocytes>10,000 | 393 | < 10,000 | 57 (14.5) | 63 (16) | 0.18 |
|  |  | > 10,000 | 110 (28) | 163 (41.5) |  |
| Lymphocytes | 387 | Menor | 165 (42.6) | 216 (55.8) | 0.66 |
|  |  | >4.00 | 2 (0.5) | 4 (1) |  |
| Neutrophils>70 | 388 | < 70 | 8 (2.1) | 7 (1.8) | 0.43 |
|  |  | > 70 | 159 (41.0) | 214 (55.2) |  |
| PaFi | 369 | >300 | 48(31) | 39(18) | **<0.001** |
|  |  | 201-300 | 52(34) | 60(28) |  |
|  |  | 101-200 | 41(26) | 87(41) |  |
|  |  | 81-100 | 6(4) | 10(5) |  |
|  |  | ≤ 80 | 8(5) | 18(8) |  |

Dyslipidemia promotes endothelial dysfunction and activation, which leads to an increase in pro-inflammatory cytokines such as interleukin 1 among others and the formation of reactive oxygen species [10, 11]. This cytokine increase could exacerbate a systemic inflammatory response in those with chronic diseases or even those who do not present them [12-14]. Once hypertriglyceridemia was defined according to the cut-off point of our laboratory as values > 150 mg/dl in blood, of the 324 patients who underwent a lipid profile in these first sixth months, we observed a higher mortality in those who presented levels > 150, being 39.2% (127) of the non-survivor’s vs 24.7% (80) of the survivors. As for those with levels <150, the observed mortality was similar in both 18.8% (61) survivors vs 17.3% (56) of non-survivors. Although we observe a higher mortality in patients with triglyceride levels > 150 mg/dl, we cannot establish a direct correlation with the level of severity of the disease.

The cytokine profile was also studied. We noted that most patients admitted to ICU (81.1%) presented high values of IL-6 (mean 248, range 1.15 -1620.0). IL-6 determination was used as an indicator to use Tocilizumab. Among those patients that received Tocilizumab, we noted that levels of IL-6 resulted higher in patients that did not survive at ICU compared to those that survived after ICU (288.4 UI vs 119.02 UI, *p*<0.001, Fisher exact t-test)

Additionally, laboratory bacterial cultures were analyzed. Nearly 386 (69.1%) showed evidence of infections associated with health care. These include 275 (49.1%) positive cultures, 183 (33%) endotracheal secretions, 55 (9.9%) urine cultures and 170 (30.4%) blood cultures. From the endotracheal secretions cultures were isolated *P. aeruginosa, A. baumannii, S. maltophilia, S. aureus, K. pneumoniae* and other species. A detailed list of isolated pathogens is shown in **Table 8**.

**Table 8:** Pathogens isolated from ICU admitted patients.

| **Endotracheal secretions** |  |
| --- | --- |
| Pseudomonas aeruginosa | 63 |
| Acinetobacter baumannii | 45 |
| Stenotrophomonas maltophilia | 37 |
| Staphylococcus aureus | 13 |
| Klebsiella pneumoniae | 10 |
| Enterobacter cloacae | 9 |
| Serratia marcescens | 5 |
| Enterobacter aerogenes | 5 |
| Pseudomonas putida | 2 |
| Pseudomonas fluorescens | 2 |
| **Blood culture** |  |
| Staphylococcus epidermidis | 182 |
| Pseudomonas aeruginosa | 124 |
| Acinetobacter baumannii | 114 |
| Enterococcus faecalis | 73 |
| Staphylococcus aureus | 71 |
| Stenotrophomonas maltophilia | 64 |
| Klebsiella pneumoniae | 53 |
| Enterobacter cloacae | 46 |
| Escherichia coli | 28 |
| Serratia marcescens | 22 |
| Enterobacter aerogenes | 19 |
| Achromobacter xylosoxidans | 12 |
| Burkholderia cepacia | 12 |
| Enterococcus faecium | 7 |
| Providencia stuartii | 6 |
| Candida tropicalis | 72 |
| Candida albicans | 41 |
| Candida glabrata | 13 |
| Candida auris | 13 |
| Candida parapsilosis | 9 |

A total of 1289 blood cultures were positive, among which 472 Staphylococcus species were identified, including *S. aureus*. Of that total, 276 (58.4) were considered contaminants. The most isolated bacteria from blood cultures were *S. epidermidis, P. aeruginosa, A. baumannii, E. faecalis, S. aureus, S. maltophilia*, detailed in Table 8. Of notable importance was the great isolation of Candida species; *C. tropicalis, C. albicans, C. glabrata* and *C. auris*.

## DISCUSSION

COVID-19 turned into the major public health threat during the 2020-2021 period. Here we aimed to describe the clinical and epidemiological characteristics of COVID-19 patients admitted to the ICU at the main COVID 19 hospital in Panama during the first pandemic wave during April to September 2020 period. We found that survival from COVID-19 is highly dependent on the presence of comorbidities and a worse biomarker profile of each patient.

High mortality rate was due to uncontrolled ARDS al ICU’s worldwide. In August 1967, Dr. David Ashbaugh, together with Boyd Bigelow, Thomas Petty, and Bernard Levin, described for the first time in 12 patients a syndrome characterized by hypoxemia, tachypnea, and decreased lung compliance that did not respond to conventional methods of oxygen therapy. The syndrome resembles that observed in childhood distress syndrome, which presents hyaline membranes and atelectasis, which is why it is called Adult Acute Respiratory Syndrome, mentioning in turn the possible beneficial impact of the use of PEEP (positive pressure at the end of expiration) and corticosteroids as adjunctive therapy in fat embolism and viral pneumonia [15, 16].

In February 1975, Robert R. Kirby et al. described the usefulness of elevated PEEP levels in acute respiratory failure as a therapeutic measure to reduce the use of oxygen at high doses and its possible adverse effects [17]. This study presenting 28 patients with multiple diseases, who were measured by parameters such as short intrapulmonary circuits (Qs / QT)., mixed venous blood (QS / QT), arteriovenous difference (CaO2-CvO2), cardiac output (CO) and PaO2 / FiO2. Cardiac output in these patients was not negatively affected at any PEEP level up to 32 torr (44 cm H2O), concluding that high PEEP levels may show therapeutic benefits for patients with refractory respiratory failure when combined with intermittent mandatory ventilation (IMV) and careful cardiovascular monitoring.

Because the patients received variable fractions of inspiratory oxygen (FI02) at different levels of PEEP, individual changes in Pa02 were standardized using the expression PaO2 / Fi02, or as it is also known today as the Kirby index, accepting PaO2 as normal levels. / FiO2> 300 mmHg [17]. Subsequent studies have shown a higher mortality the lower the index, for example, PaO2 / FiO2 <100 mmHg [18-22].

In 1988 John Murray, Michael Matthay and John Luce published an article entitled “An expansion of the definition of adult respiratory distress syndrome by establishing a scale that included oxygenation levels, PEEP, pulmonary infiltrates on chest radiography and levels of lung compliance [20].

Due to the great variability of definitions and with the purpose of unifying criteria, in 1994 the European-American Consensus Conference (AECC) was given formalizing criteria for the diagnosis of acute respiratory distress syndrome (ARDS), although this definition was simple to define apply in the clinical setting, was questioned over the years, since the evaluation of the oxygenation defect did not require standardized ventilatory support [23].

Finally, in 2012, a new definition was established in Berlin, in relation to the previous definitions, eliminating the term acute lung injury (ALI) and the pulmonary capillary pressure criterion (PCP <18), in addition to adding the ventilation settings mechanics with a minimum CPAP or PEEP setting of at least 5cm H_2_O. The authors emphasize that pulmonary edema of cardiogenic origin as well as fluid overload should be ruled out and establish the acute onset in the first week of presentation, bilateral pulmonary infiltrates on the chest radiograph, which are not explained by pleural effusion, atelectasis, pulmonary nodules, fluid overload or heart failure and finally, deterioration in oxygenation defined by the PaO2 / FiO2 ratio, which shows the degree of hypoxemia defining the severity and associated mortality [24].

ARDS is a public health problem with some geographic variations, with a high mortality being around 40% and representing 10.4% of total admissions to the intensive care unit (ICU) and 23.4% of all patients. patients who required mechanical ventilation and constituted 0.42 cases / ICU bed for 4 weeks [25].

Patients with ARDS associated with COVID-19 have a form of injury that, in many respects, is like that of those with ARDS not related to COVID-19, presenting a reduction in lung compliance and that together with an increase in D-dimer concentrations have high mortality rates.

Pathophysiologically, ARDS is characterized by acute and diffuse inflammatory damage to the capillary-alveolar barrier known as diffuse alveolar damage (DAD), which is associated with increased vascular permeability, as well as reduced compliance and tissue size. aerated lung, compromising gas exchange and causing hypoxemia. The autopsies of patients reported by Nicholls et al and Pei et al share histopathological findings of hemophagocytosis, squamous metaplasia of pneumocytes, thickening of the pulmonary septa, and intraalveolar hemorrhage [26, 27].

During the pandemic Gattinoni et al. hypothesize that the different COVID-19 patterns found depend on the interaction between the severity of the infection, the host’s response, its physiological reserve, and comorbidities; the ventilatory response capacity of the patient to hypoxemia; the time elapsed between the onset of the disease and the observation in the hospital. The interaction between these factors leads to the development of a spectrum of time-related diseases within two primary “phenotypes”: Type L, characterized by low elastance (ie, high compliance), low ventilation-perfusion ratio, low lung weight, and low recruit ability, while type H, characterized by high elasticity, high right-to-left shunt, high lung weight and high lung recruitment capacity [28].

The PaO2/FiO2 ratio was calculated for 369 patients, of which 87 patients presented values > 300 with a mortality of 18% (39 patients), with values between 300 and 200, 112 patients with a mortality of 28% (60 patients) were found. patients), in the range of 200 to 100 we found 128 patients with a mortality of 41% (87 patients), finally in the range <100, 42 patients were enrolled with a mortality of 66.7% (28 patients), which is concordant with the literature to date.

The mortality rate among COVID-19 patients who required admission to the ICU ranged from 16% to 78% [9, 29-32]. In our study, we observed a global mortality of 58.9%. The high mortality observed in our study compared to other hospitals that also handled a large volume of COVID 19 patients in other countries who included non-mechanically ventilated patients also, could be explained in part because in our study we included mostly patients intubated on invasive mechanical ventilation (96%), and if we compare this specific population (mechanically ventilated patient), the mortality observed in our study is similar to other studies with the same characteristics carried out in high-income countries as for example in the report by Docherty et al [23].

Regarding the epidemiological characteristics, our study population included nearly two thirds of men; and the age group most affected was between 56 and 75 years old similar to other studies such as those described in Seattle and Lombardy [29]. The characteristics of the patient in our study were generally like those described in patients on invasive mechanical ventilation in studies carried out in high-income countries [5, 29, 30, 33, 34]. On the other hand, we observed various comorbidities including arterial hypertension, type 2 diabetes mellitus, being related to other studies reported by Guan *et al* and Wang et al, followed by obesity, asthma and chronic kidney disease [33]. The above results confirm that non transmissible and chronic diseases are the main comorbidities highly related with COVID-19 death.

The clinical management of COVID-19 patient have been outlined. In our study, the patients received mechanical ventilation therapy according to current guidelines for the management of patients with acute respiratory distress syndrome (ARDS) by the ATS [35]. We use low Vt (tidal volume), moderate levels of PEEP (positive end-expiratory pressure), low DP (driving pressure) and low PP (plateau pressure). Such approach was like those used by others previously [6, 29]. On the other hand, several drugs were used at the beginning of the pandemic to try to save the lives of our patients based on the studies available to date including ivermectin and tocilizumab [36]. However, the medications used for COVID 19 patients were modified as new scientific evidence appeared, discarding those that the scientific evidence did not support. Thus, the clinical management based on drugs evolved together with the pandemic.

The most frequent causes of death in the ICU of the patients in our study were: Multiorgan dysfunction syndrome, followed by acute respiratory distress syndrome secondary to COVID 19 pneumonia, shock of different etiologies (septic, obstructive, cardiogenic, and mixed), acute renal failure, refractory pneumothorax complicated by bronchopleural fistula.

Blood biomarkers remained key for appropriate management of seriously ill patients with COVID-19. Overall, we observed that those patients with biomarker levels above the cut-off point (including the CRP, procalcitonin, D-dimer, creatinine, BUN, ferritin, hypertriglyceridemia, IL-6, leukocytes, lower PaO2/FiO2 ratio) had higher mortality rate. For example, emerging evidence support the notion that dyslipidemia promotes endothelial dysfunction and activation, which leads to an increase in pro-inflammatory cytokines such as interleukin 1 among others and the formation of reactive oxygen species [11, 14]. This situation could exacerbate a systemic inflammatory response in those with chronic diseases or even those who do not present them during COVID-19. In addition, we noted a higher mortality in those who presented hypertriglyceridemia as mentioned before, however, we cannot establish a direct correlation with the level of severity COVID-19 and causality. Altogether, the blood biomarkers remain as main clues to predict mortality outcome among ICU admitted patients.

Another cause of early death among COVID-19 patient admitted to ICU includes infectious complications. As their stay is prolonged, nosocomial infections complication arise, complicating the condition and inducing mortality. In our study, nearly three quarters of the patients had an infection associated with clinical management at hospital. Similarly, all other changes during pandemic peak increases like in different countries in this first sixth months including: 1) ICU room remodeling; 2) rushed training for non-ICU personnel; 3) lack of personal protection implements; and 4) fear of personnel to handle COVID-19 patients. This situation become complicated with nosocomial infection with multidrug resistant strains [37]. All together, these factors contributed to the mortality rate observed in our study. Further efforts are warranted to assure clinical management of critical patient, especially during crisis conditions such as the current pandemic.

Multidrug resistant bacteria complicate the management of critically ill COVID-19 patients [37]. In our study, we observed *Pseudomonas aeruginosa, Stenotrophomonas maltophilia* and *Acinetobacter baumannii* predominated in cultures of endotracheal secretions. Regarding blood cultures, *Staphylococcus* species predominated, interpreted mostly as contaminants, indicating the difficulty in obtaining good blood samples in these patients due in part to the personal protective equipment that health personnel must wear. The same bacteria found in endotracheal secretions predominated as blood pathogens. Exception made in the finding of *Enterococcus faecalis*, which may indicate the difficulty in cleaning these patients. The large number of blood isolates of Candida species is striking, this because of the massive use of antimicrobials for these patients. The appearance of *Candida auris*, a new emerging species, represents the persistence of this yeast after several outbreaks that have occurred in this hospital. In summary, the management of COVID-19 patient admitted in ICU requires higher effort to avoid the transmission of drug resistant bacteria to favor survival.

The sustained burden on health personnel by COVID-19 could also have contributed to the high mortality in Panama observed in the first six months of the pandemic.

This study has several limitations. First, among the limitations we highlight those inherent to the type of study carried out (cross section descriptive observational study). We did not study causality, we only limited ourselves to describing our population.

Second, the nature of the database did not allow more detailed information to be obtained, such as ventilatory monitoring of days after baseline or more specific laboratory data taken on days other than those officially designated for the study (for example, on weekends, no data was recorded in this regard). The number of cases is small, so there may be independent determinants of mortality that could not be identified. The sustained work burden on health personnel by COVID-19 could also have contributed to lack of some important information on medical records on the specific days designated for obtain it. Thus, further studies should allocate dedicated resource to tackle these limitations and assure a complete data set.

## CONCLUSIONS

COVID-19 mortality rate upon admission to ICU in the first sixth month of pandemic rely on epidemiological and clinical patient characteristics, blood biomarkers and clinical management. The survival of critically ill COVID-19 patients greatly depends on an accurate management based on the interaction with all above factors. Our study provides the most common characteristics of these patients, including most of clinical variables similar to those described to date in other international reports. Such resource will aid in being able to better understand and set up the best strategy to save lives of the critical ill COVID-19 patients. Future studies should focus efforts to understand causality effect of every risk factor on inducing death after admission to ICU. Such strategy will provide concepts of evidence-based medicine and thus be able to contribute to create more appropriate management strategies for COVID-19 patients. Until now, our study only provides the baseline characteristic of ICU admitted patient. We believe, above evidence will help on the global effort on developing promising medications to treat severe COVID-19. In the meantime, supportive care continues to be the cornerstone for its management, ventilation strategies, hemodynamic control, fluid administration and prevention of thromboembolic complications (with anticoagulants), corticosteroids, prevention, and treatment of coinfections.

## Data Availability

All data produced in the present study are available upon reasonable request to the authors

## ACKNOWLEDGEMENTS

We thank all the patients affected by COVID-19 for their participation in our study. We also thank the health care workers from all ICU at Complejo Hospitalario Metropolitano Dr. Arnulfo Arias Madrid for their contributions to patient information and providing access to patient’s files. Lastly, we thank Colleen Goodridge for her critical review of the manuscript and valuable suggestions.

